# Leveraging Machine Learning & Mobile Application Technology for Vitiligo Management: A Proof-of-Concept

**DOI:** 10.1101/2024.09.06.24313068

**Authors:** Mahla Abdolahnejad, Hyerin Jeong, Victoria Lin, Tiffany Ng, Emad Altaki, Anthea Mo, Burak Yildiz, Hannah O. Chan, Collin Hong, Rakesh Joshi

## Abstract

Vitiligo, a dermatological condition characterized by depigmented patches on the skin, affects up to 2% of the global population. Its management is complex, often hindered by delayed diagnosis due to limited access to dermatologists and/ digital tools. Recent advancements in machine learning (ML) offer a potential solution by providing digital tools for early detection and management. This proof-of-concept study describes the development of a machine learning pipeline integrated into a mobile application for vitiligo assessment.

Using a dataset of 1,309 images, including segmental and generalized vitiligo, the CNN was trained for binary classification with an accuracy of 95%. The model segments depigmented patches and conducts colorimetric analysis for precise evaluation. We compared traditional Wood’s lamp imaging with CNN-generated maps, showing comparable or superior results in detecting faint depigmentation.

Developed using Flutter for cross-platform compatibility, the app enables patients to upload images for analysis and track disease progression. A Golang-based backend ensures robust data management, while a PostgreSQL database supports secure storage of patient information. The integration of Azure Active Directory enhances security and user authentication.

This approach aims to bridge the gap in dermatological care by providing an accessible, ML-driven solution for vitiligo management. Future iterations will expand the applications’ capability to screen for other depigmentation disorders, incorporate automated scoring systems for more personalized patient management, and communication services.

## Introduction

Vitiligo is a dermatological condition characterized by the loss of melanocytes resulting in depigmented patches on the skin [1,2]. The condition affects 0.5-2% of the global population [2,3]. Although the exact cause of vitiligo remains unclear, it is believed to be the result of a combination of autoimmune, metabolism, oxidative stress, genetic, and environmental factors [3]. Clinically, vitiligo presents as well-demarcated, depigmented macules and patches, often symmetrical, that can appear on any part of the body. The presentation can be segmental (on one side of the body) or generalized [1,4].

Diagnosing vitiligo is primarily based on clinical observation, supplemented by tools like Wood’s lamp examination, dermatoscopy, and histopathology when necessary [1,3,5,11]. However, there is no single definitive diagnostic test, making the role of experienced dermatologists crucial in distinguishing vitiligo from other hypopigmented conditions, like Tinea versicolour, a fungal infection [1,5,6]. Unfortunately, access to dermatologists, is limited in many regions, which leads to delayed diagnosis and treatment [3].

Vitiligo can profoundly impact the patients’ quality of life, both physically and psychologically [7,8]. Recent studies have highlighted the substantial emotional and social burden associated with vitiligo, including patient-centric grass root movements to improve the quality of life for vitiligo patients [9]. These studies underscored the psychological impact of vitiligo and emphasized the importance of timely, accurate diagnosis and management. It also highlights the need for access to mobile digital tools that can be used by patients to not only self-assess and track their diseases, but also provide education, support, and access to medical professionals [10,11].

The integration of machine learning (ML) into dermatology has opened new avenues for improving screening [12]. Artificial Intelligence models, particularly deep learning networks, have shown significant potential in assisting with the management of various skin conditions, including skin lesions and burn injuries [12-15]. These models are capable of processing and learning from large volumes of image data, identifying patterns and features that may be subtle or overlooked by human examiners, thereby aiding in early diagnosis and personalized treatment planning.

In addition to classification tasks, recent advancements in ML have focused on improving model interpretability, especially for clinical segmentation tasks, which is crucial for clinical applications. Saliency maps like Class Activation Maps (CAMs) provide visual explanations of the model’s decision-making process by highlighting the most informative regions in the input images [16]. Abdolahnejad et al. (2023) have made significant contributions to this area through the development of Boundary Attention Mapping (BAM), a type of saliency maps for segmentation and attention mapping, for a skin use case (burn injuries). BAM is a technique that refines the attention mechanisms, using Gradient-weighted Class Activation Mapping (Grad-CAM), and early Activation Channel maps, from Convolutional Neural Networks (CNN), to accurately delineate skin abnormality boundaries in mobile device-captured images. These advancements have also enhanced the transparency and reliability of AI models in dermatology and surgery, making them more suitable for clinical use [14-15].

Previous research has explored various neural network architectures in the context of vitiligo assessments [17-19]. A study by Guo et al. (2022;17) presents a novel deep learning-based hybrid AI model designed to detect and assess the severity of vitiligo lesions. The model integrates the YOLO v3 architecture for effective depigmentation localization with a UNet++ architecture for detailed segmentation. Trained predominantly on images from individuals with Fitzpatrick skin types III and IV, the model achieved an accuracy of 89.7% and an Area Under the Curve (AUC) of 0.91. The results showed a strong alignment with evaluations by dermatologists, underscoring the model’s potential utility. However, the system had various limitations that can impact negatively in clinical settings or remote patient usage for vitiligo assessment, including segmentation accuracy of the UNet++ model, and its ability to generalize due to training data of a small range on skin tone spectrum, as was seen when tested on images with there is un-even lighting.

Another study [18] evaluated the performance and interpretability of deep learning models, specifically ResNet and Swin Transformer architectures, for the diagnosis of vitiligo from dermatoscopic images. The research involved a comparative analysis of five models—ResNet34, ResNet50, ResNet101, Swin Transformer Base, and Swin Transformer Large—under uniform conditions to identify the best-performing model. The Swin Transformer Large model emerged as the most effective, achieving an accuracy of 93.82% and an AUC of 0.94, with a sensitivity of 94.02% and specificity of 93.5%. The study also incorporated class activation mapping (CAM) to enhance the interpretability of the model by visually highlighting the regions in the images that contributed to the diagnosis. Here too the authors highlighted some limitations, including generizability and the effectiveness of segmentation even when using CAM.

Nevertheless, the small-scale study by Hillmer et al [19], conducted using images of facial vitiligo and a trained CNN, shows that ML models can attain at par competencies with clinicians.

Even though these studies highlight the potential of using ML models in vitiligo assessments, they fail in real-world usability as the models are not incorporated into digital tools like mobile applications with backend support for patients to use, like that described by Nugraha et al [10]. Building medical software around these ML pipelines is imperative. However, it must be built with a user-friendly design, without compromising responsiveness, reliability, and technical robustness, both in the front-end user interface software, and the backend support architecture [20].

This current study builds upon these ML and digital tool advancements, by proposing a ML-based approach that utilizes CNNs and accompanying algorithms, which has been fine-tuned for vitiligo assessment using images captured from both dermatoscopic and mobile sources.

This approach seeks to address the disparity in access to expert dermatological care by providing a digital assessment tool that can be widely deployed as a mobile application, thus improving vitiligo assessment access, tracking, and ultimately patient outcomes This paper details the methodology, preliminary results, and software framework for deploying a ML pipeline for vitiligo as a proof-of-concept, patient -acing mobile application. The application currently allows user profile creation, repository for medical history, and computer-vision based segmenting, and colorimetric analyses of de-pigmentation.

## METHODS

### 1. Data Preprocessing and Augmentation

The initial step in preparing the dataset for training a Convolutional Neural Network (CNN) involved preprocessing open-source and proprietary, patient-consented images of vitiligo and normal skin. The dataset (1309 images) consisted of 474 images of segmental and generalized vitiligo, and 835 images of normal skin. Images were resized to 600 × 600 pixels and normalized to ensure uniformity across the dataset. Data augmentation using transformation techniques such as rotation, flipping, and shifts were applied to increase the diversity of training data and enhance model robustness against variations in image capture.

### 2. Training the Convolutional Neural Network (CNN)

The CNN, built using EfficientNet B7 architecture, was trained for binary classification to distinguish vitiligo from normal skin. A K-fold cross validation method was used to split the data, with K=5. The CNN used a categorical cross-entropy loss function and the Adam optimizer, with a learning rate of 1.e^-6^. Regularization using dropout rates (0.3 to 0.5) and batch normalization were implemented to reduce overfitting. The classes were weighted (1, 1.75) to handle the imbalance in images between the two classes. The model was trained for a 20 epochs with a batch size of 4. Up to 90 layers were unfrozen during the final training round.

### 3. Segmentation Identifying Vitiligo Visual Attributes

The trained CNN binary model was integrated into a pipeline, which identifies images with vitiligo present. The pipeline begins with image preprocessing to remove of non-skin parts (such as background) from the input image. Once preprocessed, the image is passed through the trained CNN. If the CNN detects vitiligo in the image, activation maps are then extracted from the model’s first convolutional layer. The activation maps extracted from the CNN provide valuable information by highlighting specific regions of the image that display specific attributes. These maps can function as saliency maps, drawing attention to areas of interest within the image. By selecting the specific map that best highlights vitiligo attributes, we can accurately identify and analyze vitiligo patches in the color images. The identified vitiligo patches are then segmented using K-means clustering for further colorimetric analysis. The segmentation is enhanced using the Boundary Attention Mapping (BAM; Abdolahnejad et al, 2023), which improves the precision of boundary detection around the vitiligo patches.

### 4. Wood’s Lamp Comparison & Colorimetry

Images were collected from three patients diagnosed with various degrees of generalized vitiligo at a dermatology clinic. The patients provided informed consent prior to image capture. Multiple images of affected skin areas were captured using an iPhone, which provided high-resolution images under ambient lighting conditions. In addition, the same areas were photographed under a Wood’s lamp, a specialized ultraviolet light that enhances the contrast of depigmented areas, making vitiligo patches more distinct.

To evaluate the effectiveness of Wood’s lamp imaging in enhancing the detection of vitiligo, versus the saliency maps and segmentation masks generated from 2D colour images, they were aligned and compared to each other. Clinicians visually inspected and compared these paired images to assess the differences in the visibility and delineation of vitiligo patches by the two methods.

The segmented patches from the activation and BAM maps were subsequently analyzed using colorimetric analysis. This process involved examining the color image pixels within each segmented cluster to quantify the extent and severity of depigmentation. By analyzing these color pixels, grouped into distinct clusters, we achieved a more precise and detailed assessment of vitiligo severity. The results of this analysis include area percentages and average color values, reported in hexadecimal format.

### 5. Development of a Flutter-based Mobile Application

The vitiligo assessment mobile application was developed using the Flutter framework. The design focused on simplicity and accessibility, ensuring intuitive navigation for users tracking their de-pigmentation condition. Secure user authentication was implemented using Azure Active Directory (Azure AD). To enhance user engagement and data collection, a dynamic questionnaire feature was developed within the app. Utilizing Flutter forms, the questionnaire allowed users to input detailed information about their skin de-pigmentation, symptoms, and other relevant health factors.

The core functionality of the app involved the integration of a trained AI model based on the EfficientNet B7 architecture. This model was deployed as a backend service, analyzing user-submitted images and data inputs. The app communicated with the AI model via RESTful API calls, sending images (captured by the user) and receiving analysis results. These results, including classification, segmentation data and colorimetric, were then displayed to the user through the app’s interface, providing personalized feedback on their condition. Continuous integration and deployment (CI/CD) pipelines were established using GitHub Actions.

### 7. Backend Development for the Vitiligo Assessment Application

The backend was built using Golang (Go) with the Gin framework. For data management, a PostgreSQL database was implemented. The database was used to store, update, and retrieve user data, including images, health information, and AI analysis results. Atomic transactions ensured that all operations were completed successfully before committing changes, thus maintaining data consistency and integrity even in high-concurrency environments. To manage deployment and scaling, each component of the backend, including the Golang server, AI models, and the database, was containerized using Docker. These containers were then deployed into a Kubernetes cluster, which provided an orchestration layer to manage these containers efficiently. The backend followed a cloud-native architecture, ensuring that each microservice could be deployed across different cloud providers. This entire backend system is a part of an “Operating system” platform, a smart Electronic Medical Record (EMR) system, which provides clinical connectivity between the user-facing mobile application and clinicians.

## RESULTS

### Artificial Intelligence pipeline

Figure 1 illustrates the ML pipeline developed to assess the visual attributes of vitiligo from images captured using either a dermatoscope or a mobile camera. The pipeline is designed to provide a preliminary assessment of vitiligo characteristics, namely depigmentation patch segmentation and a colorimetric analysis. These images serve as the primary data source for the vision component of the pipeline. A processing algorithm is also used at this early stage to remove non-skin background of the image (Figure1 A).

**Figure 1:**
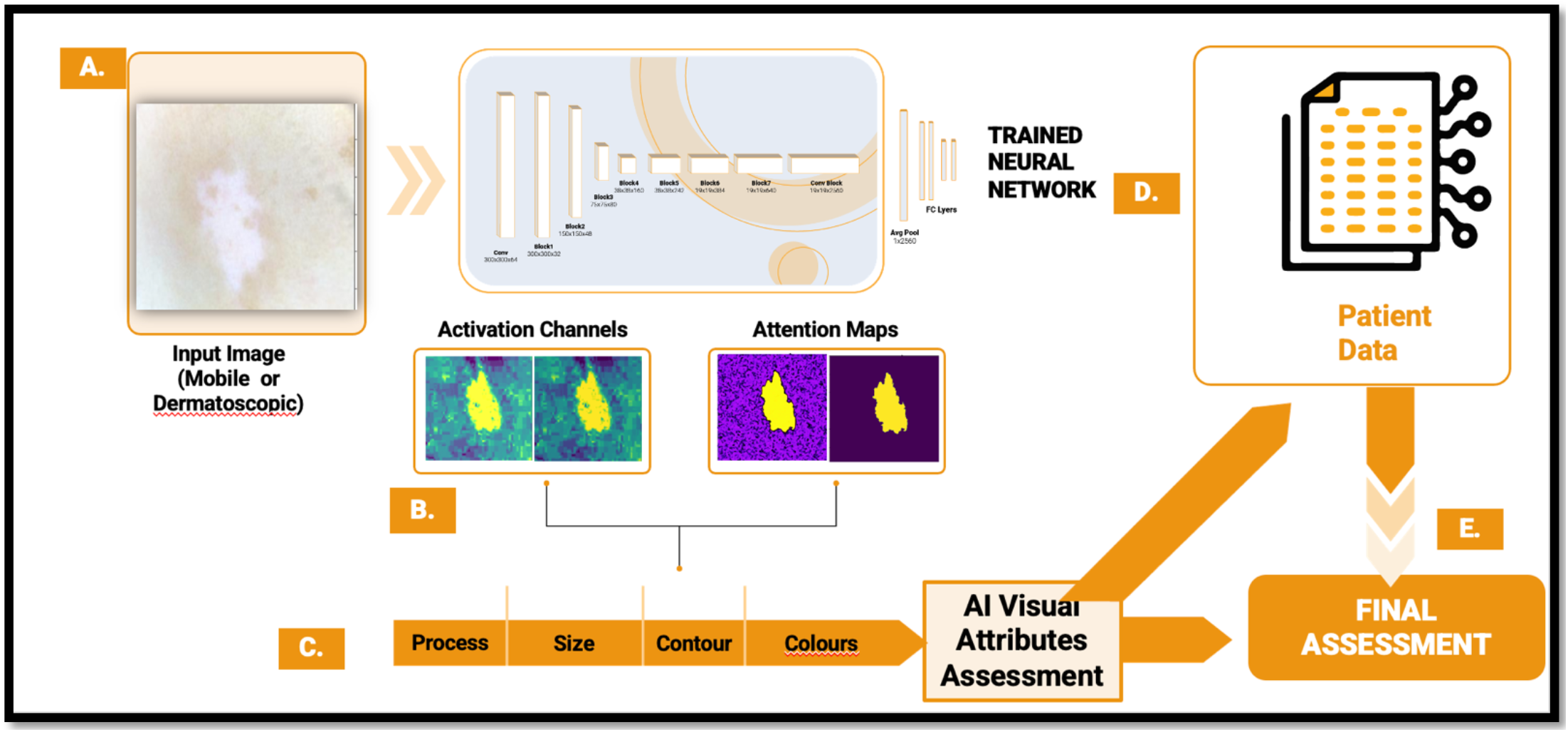
Vitiligo Machine Learning Pipeline. **A)** The schema outlines the ML-driven process for assessing de-pigmentation patches using input images. The system accepts images captured via mobile devices or dermatoscopic equipment, which are then processed through a trained neural network. **B)** The neural network generates activation channels and attention maps that highlight critical features. **C)** After map processing, size (if a fiducial marker is detected), contour, and color of the patch are detected. **D)** These visual attributes <u>can be combined</u> with patient data to provide clinicians to deliver a comprehensive final assessment (**E**) regarding the progression or status of de-pigmentation.

Next, the model extracts crucial visual features through activation channels and generates attention maps, including specialized Boundary Attention Maps (BAM; Abdolahnejad et al, 2023). The Activation channel maps highlight areas within the image that the model identifies in terms of colours and features like texture. The attention and BAM maps further refine this by indicating the specific regions where the model focuses its analysis, such as the depigmented patches characteristic of vitiligo (Figure 2 B). This visual information is used in edge and segmentation algorithms to find the size (if a fiducial marker is present in the image), the contour derived from the segmentation, and colorimetry (Figure 1C).

**Figure 2:**
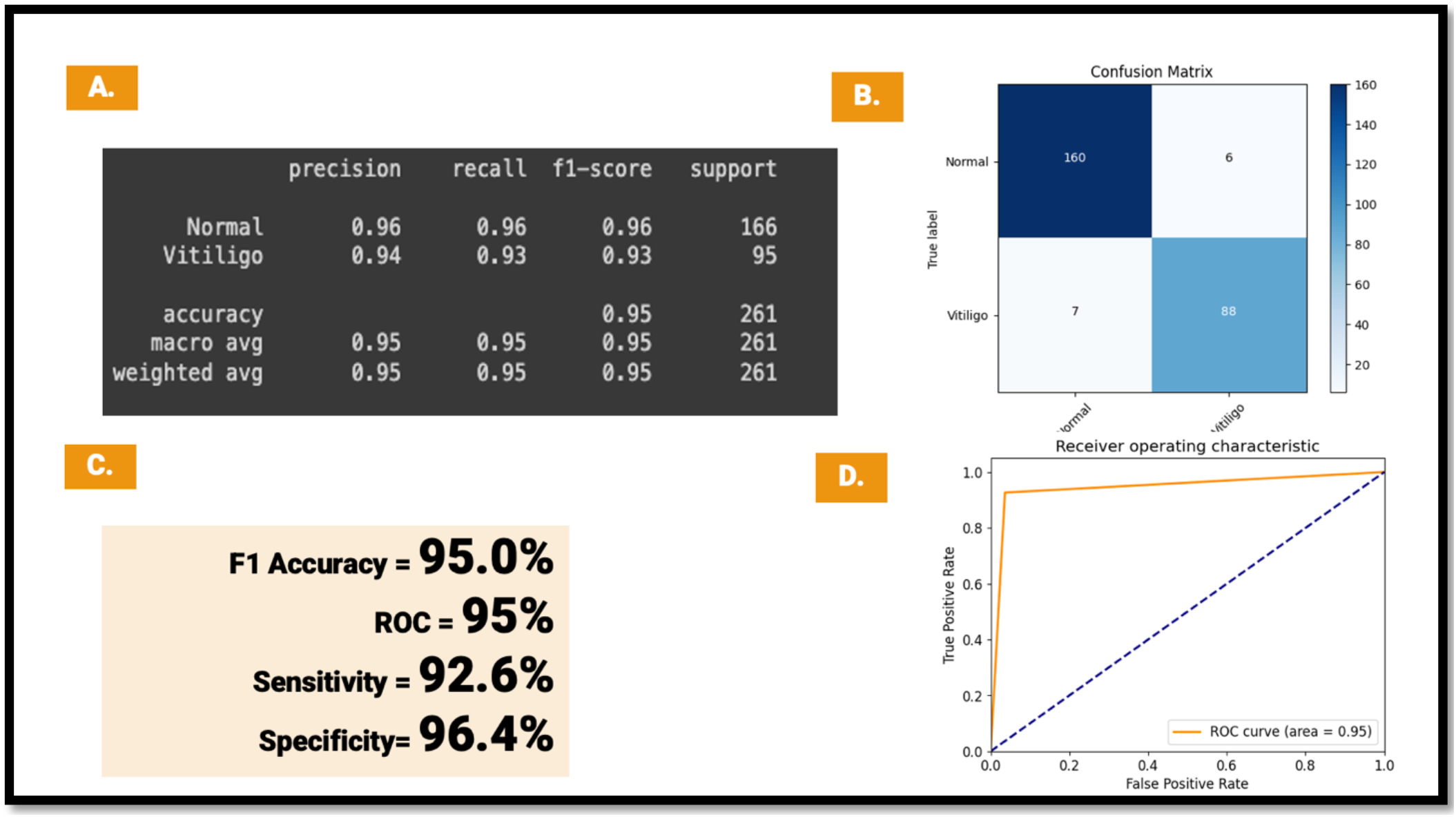
Convolutional Network Performance Metrics. **A**) Classification report summarizing precision, recall, F1-score, and support for both classes (Normal and Vitiligo), with the model achieving an overall accuracy of 95%. **B**) The Confusion Matrix illustrates the distribution of true positive, true negative, false positive, and false negative predictions, with the model correctly identifying 160 Normal cases(out of 166) and 88 Vitiligo cases(out of 95). **C**) Summary of key CNN performance metrics, including F1 Accuracy (95.0%), ROC (95%), Sensitivity (92.6%), and Specificity (96.4%), indicating high model performance in discriminating between the two classes. **D**) Illustration of the Receiver Operating Characteristic (ROC) curve, with an area under the curve (AUC) of 0.95, further indicating the model’s strong discriminative ability between Normal and Vitiligo cases.

Once visual information has been extracted, then patient data is integrated into the analysis. This data includes self-identified demographic information, medical history, and other relevant health questions that can influence the clinical assessment of vitiligo (Figure 1D).

The integration of patient data with the AI-driven visual attributes improves the relevance of the data that a trained medical professional requires to make a definitive assessment. The comprehensive visual and patient data analysis allows for a more informed diagnosis and management plan for patients with vitiligo, aiding clinicians in tailoring treatments based on the precise characteristics of the condition observed in each patient (Figure 1E).

### Convolutional Neural Network Metrics

The key performance metrics of the CNN are presented in Figure 2. The model attained a harmonized F1 accuracy of 95.0%, indicating a high balance between precision and recall in its classifications, even though there was a 1:1.75 imbalance between the classes (Figure 2A). There were signs of initial overfitting that was corrected by increasing the drop rates and lowering the learning rate for the Adam optimizer. The model also achieved a true positive rate (sensitivity) of 92.6% and a true negative rate (specificity) of 96.4%. This is supported by the provided confusion matric, further emphasizing its reasonable performance in correctly identifying both positive and negative cases (Figures 2 B, C). The Receiver Operating Characteristic (ROC) curve exhibited an area under the curve (AUC) of 95%, confirming discriminative ability between the two classes—vitiligo and normal/healthy skin (Figure 2D).

### Segmentation, Wood’s Lamp Comparison & Colorimetry

Clinicians noted that Wood’s lamp images provided of the 3 patients (Figure 3. Panel I), show clearer boundaries of depigmentation compared to ambient light 2D colour images (Figure 3, panel II). This confirms that Wood’s lamp UV imaging is a valuable tool in clinical settings for enhancing the detection and assessment of vitiligo patches. However, a qualitative analysis indicates that the CNN generated maps, both activation maps and masks (Figure3, panels III and IV, respectively), provided at-par results (Figure 3, panel B), or improvements for detecting fainter patches, or delineating the boundaries even when de-pigmentation was faint (Figure 3, panels A, C). Figure 4A depicts an example of the resultant segmentation of a colour image with a patient with vitiligo. The K-means clustering algorithm for colorimetry provides colours detected as percentage values and tags the colour with a hexadecimal value for standardization, which will facilitate for the tracking of changes in colour composition of de-pigmented skin patches over time.

**Figure 3:**
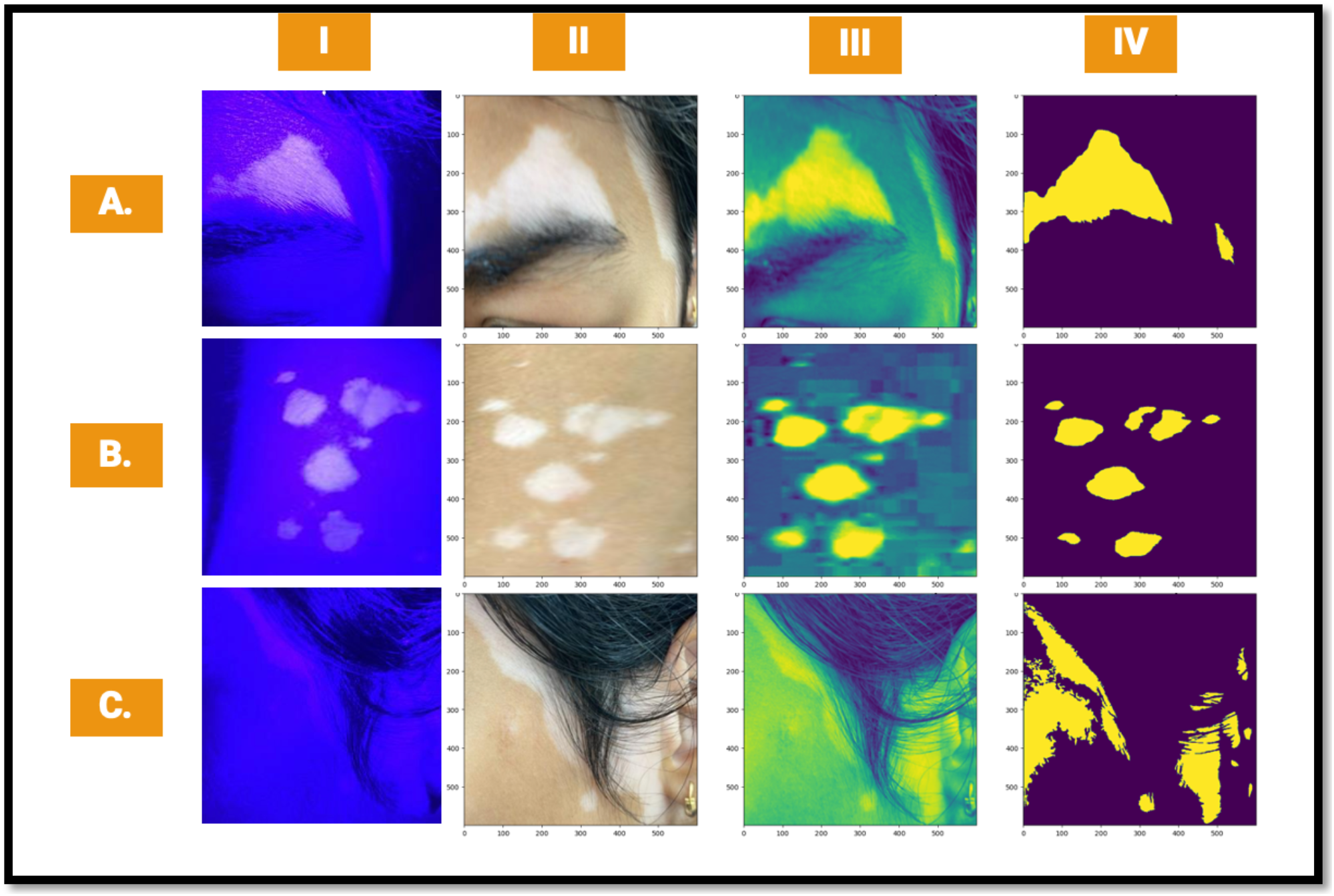
Wood’s Lamp Comparison. This figure compares Wood lamp images (UV, **Panel I**) with corresponding 2D color images (**Panel II**), CNN-derived activation maps(**Panel III**), and final segmentation masks(**Panel IV**) across three patients with vitiligo (**Panels A, B, and C**). **Panel I** (UV Wood Lamp Images) depicts vitiligo patches under UV light for three different patients, highlighting depigmented areas. **Panel II** (2D Color Images) illustrates the same vitiligo patches under normal light for each patient, showing the visible appearance of the patches. **Panel III** (CNN Activation Maps) displays the activation channels generated by the CNN, highlighting key areas of interest for patch detection and analysis. **Panel IV** (Final Masks provides the final segmentation masks of vitiligo patches derived from the BAM system, isolating depigmented regions for precise quantification and analysis. The horizontal panels (**A, B**, and **C**) represent three different patients with vitiligo, demonstrating consistent detection across various imaging modalities.

**Figure 4:**
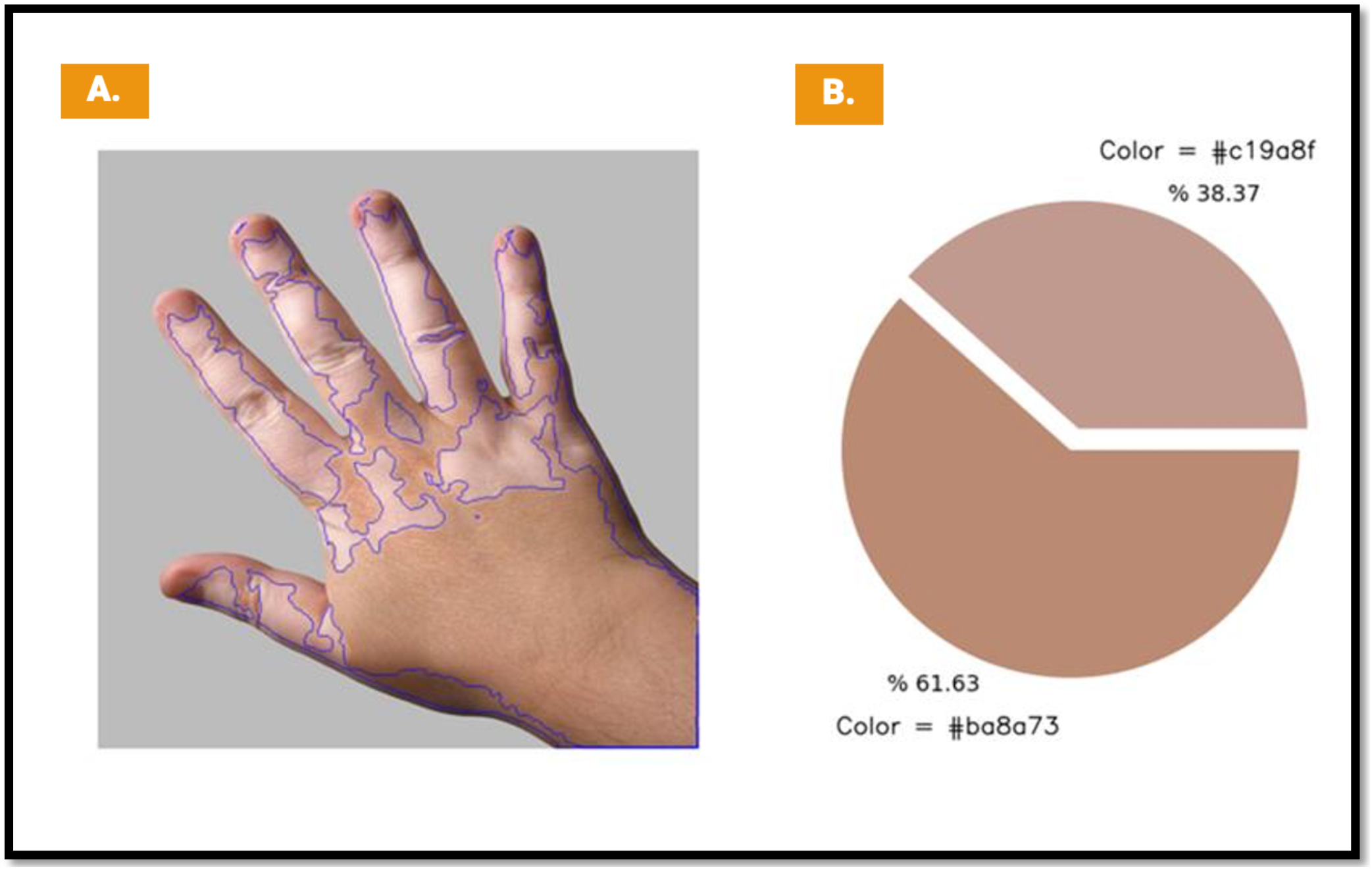
Segmentation & Colorimetric outputs for Mobile Application users. Outputs utilizing the vitiligo machine learning pipeline includes contour mapping and color analysis. **A)** Display de-pigmentation on a hand, where the application has identified and outlined individual depigmented areas (*in blue*). These contours allow visual tracking of patch boundaries and changes over time by comparing time-series images. **B**) A pie chart breaks down the color composition of the hand, providing percentages of different skin tones. The application detects two dominant skin colors, assigning hexadecimal codes and quantifying the area covered by each (% 38.37 and % 61.63), which helps in monitoring depigmentation (***NB***. The current version of the Application does not include patch dimensions in this output, the embedded algorithm does have that capability, with the use of fiducial markers).

### Frontend & Backend Development

To deploy the Vitiligo AI pipeline, a Flutter software framework was chosen for its capability to create high-performance, cross-platform applications from a single codebase, enabling deployment on both iOS and Android devices. Figure 5 shows examples of the Mobile Application pages, for secure login (Figure 5 A), photo upload for AI analysis (Figure 5 B), Questionnaire for capturing patient data like medical history (Figure 5C), and an AI results page (Figure 5 D).

**Figure 5:**
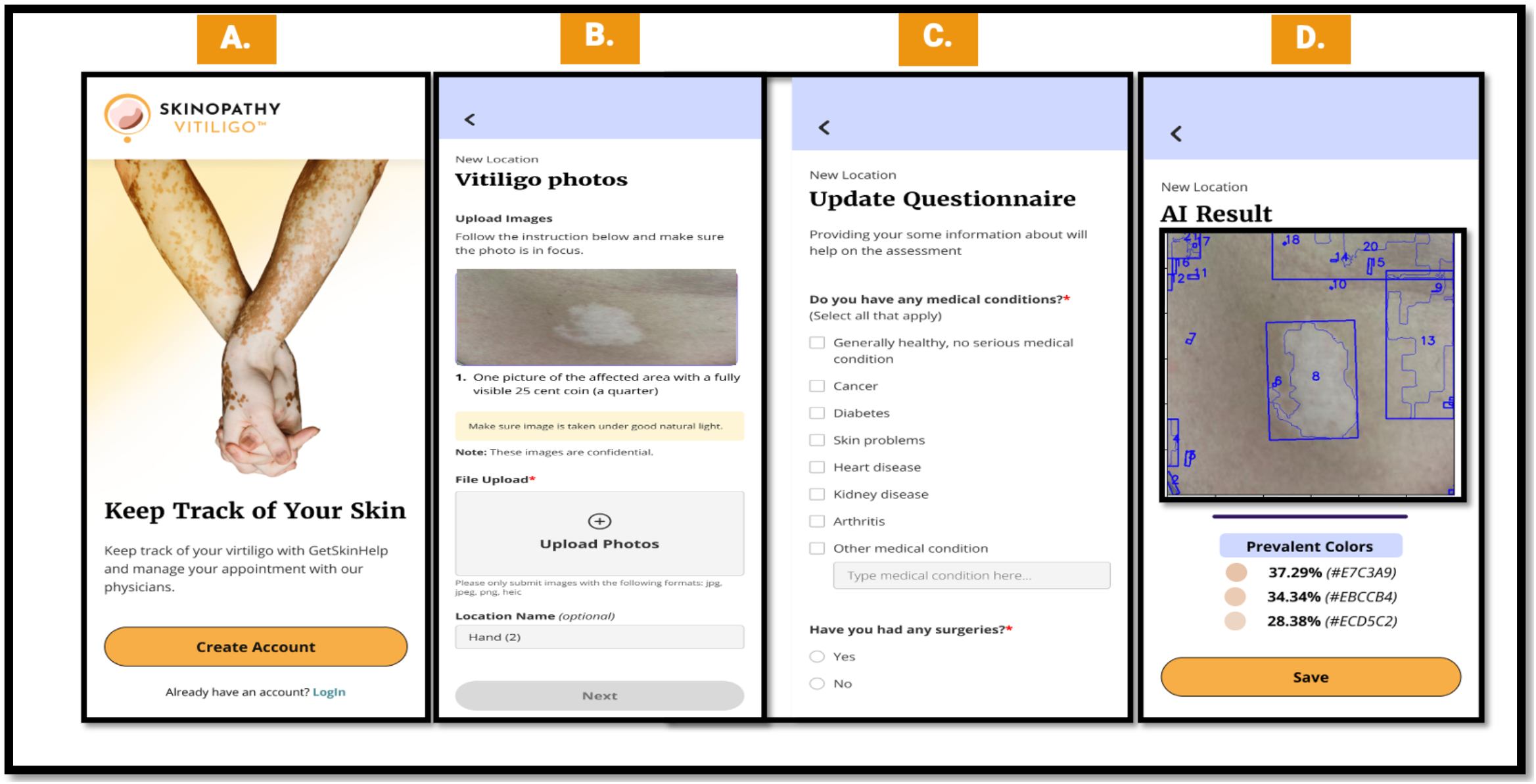
A Mobile Applications for Vitiligo Patients: The user interface of the Vitiligo mobile application, guides users through the process of vitiligo tracking and AI-based assessments. A) The home screen page displays the app’s welcome page, encouraging users to create an account to monitor their vitiligo progression. It offers them a link to use a virtual medical service where they can get appointments with a medical professional **B**) The Photo Upload page prompts the user to upload images of their depigmented areas. The instructions specify the inclusion of a coin in the image (for reference size) to aid in estimating the size by the user and clinicians **C**) A Medical Questionnaire page encourages users to fill out relevant health information, such as any existing conditions (e.g., diabetes, heart disease) and history of surgeries, to personalize the assessment and assist in understanding the progression of vitiligo by clinicians. **D**) An AI results page displays the AI-generated result (See Figure 4 for details). The application can also include pages for patient consent for several data-related activities: consent for use of de-identified data for research and development, consent to be contacted to participate in any future Vitiligo study, consent for notification of new products for disease management and support group information, etc. (*pages not shown)*.

Azure AD integration of the mobile application provided robust identity management and access control, ensuring that user data, particularly sensitive health information, was securely handled, as was seen by internal development environment testing. The patient questionnaire was designed to be adaptive, adjusting its questions based on user responses to capture personalized data that can be used for a clinical diagnosis when all the data from the app is provided to a health provider. Continuous integration and deployment (CI/CD) pipelines were established using GitHub Actions to maintain the app’s reliability and ensure timely updates.

For the backend, a Go (Golang) framework was selected for its advantages in terms of response speed, computational efficiency, and its ability to handle high concurrency, making it ideal for applications requiring real-time processing and quick responses. The Gin framework provided a lightweight and fast web framework, facilitating the development of RESTful APIs that efficiently handled user requests and communicated with the AI pipeline’s inference services.

PostgreSQL was chosen for its robustness as a relational database system, offering dedicated support for complex queries, data integrity, and atomic transactions. Kubernetes allowed the application to handle demand spikes by scaling distinct parts of the backend independently, ensuring that resources were allocated dynamically based on current needs. This setup also enabled the backend to achieve high availability and fault tolerance. This architecture allows the services to communicate with each other in a fault-tolerant manner, handle failures gracefully, and self-heal through replicated deployments. The cloud-native approach also support<u>s</u> continuous delivery and scalability, which are critical for maintaining the application’s performance and reliability as user demand grows. As this backend is a part of a smart EMR platform, this architecture allows for connectivity between the user-facing Mobile Application and the Clinician-facing OS platform. If a user consents to become a patient, this integrated system can allow for physicians to see the user as a patient and provide medical services.

## Conclusions & Future Work

Overall, the results indicated that the EfficientNet B7-based CNN is effective in classifying vitiligo images, making it a reliable tool for aiding in the preliminary assessment of vitiligo from images. The combination of high accuracy, strong ROC performance, and balanced sensitivity and specificity suggests that this model can be used with some confidence in helping segment depigmented patches and in aiding colorimetry analysis. A future iteration of the CNN will be to train the model on vitiligo, healthy skin as well as images of other de-pigmentation diseases.

This will allow the system to conduct an initial screen of whether the de-pigmentation is due to vitiligo or some other etiology. The UV to AI image comparisons also provides convincing evidence for further investigation. A large-scale study would validate the Vitiligo AI pipeline’s potential to replace Wood’s Lamp analyses when one is not available.

Further additions to the AI pipeline can include automated VASI scoring using a hybrid computer vision and questionnaire system. This would demonstrate the efficacy of an ML pipeline in providing a nuanced and data-driven assessment of vitiligo, leveraging both advanced image processing techniques and patient-specific information to deliver personalized and accurate evaluations. Providing treatment options for clinicians who can see the patient data and AI results, through the <u>M</u>obile <u>A</u>pplication-OS integration, would also help manage vitiligo.

By using Flutter, Golang, PostgreSQL, Docker, Kubernetes, and cloud-native principles, the Mobile Application front-end and the backend are equipped to handle the complex demands of the vitiligo assessment application, providing a reliable foundation for the AI-driven services, and ensuring seamless operation under varying load conditions. Using these Frontend/Backend system<u>s</u>, also allows the integration of communication services and virtual health portals, for patients to engage in virtual consultations for their disease status. These advances will allow remote and asynchronous monitoring of vitiligo patients, provide motivation for the patient to adhere to treatments by allowing tracking of the condition by the patient, and reduce the burden on clinicians.

## Data Availability

All data produced in the present work are contained in the manuscript

